# Non-invasive detection of bilirubin concentrations during the first week of life in a low-resource setting along the Thailand-Myanmar border

**DOI:** 10.1101/2024.05.06.24306917

**Authors:** Germana Bancone, Mary Ellen Gilder, Elsie Win, Gornpan Gornsawun, Paw Khu Moo, Laypaw Archasuksan, Nan San Wai, Sylverine Win, Borimas Hanboonkunupakarn, Francois Nosten, Verena I Carrara, Rose McGready

**Affiliations:** Shoklo Malaria Research Unit, Mahidol-Oxford Tropical Medicine Research Unit, Faculty of Tropical Medicine, Mahidol University, Mae Sot, Thailand; Centre for Tropical Medicine and Global Health, Nuffield Department of Medicine, University of Oxford, Oxford, UK; Mahidol-Oxford Tropical Medicine Research Unit (MORU), Faculty of Tropical Medicine, Mahidol University, Bangkok, Thailand; Institute of Global Health, Faculty of Medicine, University of Geneva, Geneva, Switzerland

## Abstract

**Background:** Neonatal hyperbilirubinaemia (NH) is a common problem worldwide and causes morbidity and mortality especially in low-resource settings.

**Methods:** A study was carried out at Shoklo Malaria Research Unit clinics along the Thailand-Myanmar border to evaluate a non-invasive test for diagnosis of NH in a low-resource setting. Performance of transcutaneous bilirubinometer Dräger Jaundice Meter JM-105 was assessed against routine capillary serum bilirubin testing before phototherapy during neonatal care in the first week of life. Results were analysed by direct agreement and by various bilirubin thresholds used in clinical practice. Total serum bilirubin was also measured in cord blood at birth and tested for prediction of hyperbilirubinaemia requiring phototherapy in the first week of life.

**Results:** Between April 2020 and May 2023, 742 neonates born at SMRU facilities were included in the study. A total of 695 neonates provided 1 to 9 capillary blood samples for analysis of serum bilirubin (total 1244 tests) during the first week of life and performance of the transcutaneous bilirubinometer was assessed in 307 neonates who provided 687 paired transcutaneous-capillary blood tests. Bilirubin levels were also measured in 738 cord blood samples.

Adjusted values of the transcutaneous bilirubinometer showed excellent agreement with capillary serum bilirubin concentration (intraclass correlation coefficient=0.923) and high sensitivity (>98%) at all clinical thresholds analysed across three years of sampling and multiple users. Concentrations of bilirubin detected in cord blood were not useful in identifying neonates at risk of hyperbilirubinaemia requiring treatment.

**Conclusions:** The transcutaneous bilirubinometer is a reliable tool to screen neonates and identify those needing confirmatory blood testing. Bilirubin concentrations in cord blood are not predictive of hyperbilirubinemia in neonates.

**Summary box:** *What is already known on this topic:* Non-invasive detection of bilirubin levels in cord blood and transcutaneously can support better clinical care of neonates at risk of hyperbilirubinaemia, especially in low resources settings.

*What this study adds:* This study was the first carried out in neonates of Karen and Burman ethnicity born at the Thailand-Myanmar border. The study provides new data on the performance of a transcutaneous bilirubinometer used by locally trained birth attendants. The results show that cord blood bilirubin levels are not predictive of hyperbilirubinaemia risk in the first days of life.

*How this study might affect research, practice or policy:* This study adds to the growing body of knowledge about performance and utility of non-invasive screening tools and diagnostics to improve neonatal health in low-resource settings and LMIC countries.

## Introduction

Physiologic jaundice is common in newborns and usually resolves spontaneously within two weeks. However, when unconjugated bilirubin blood exceeds an age-dependent concentration (resulting in neonatal hyperbilirubinaemia, NH), it can pass the blood-brain barrier and lead to serious complications, including neurologic damage and death.

Several risk factors are associated with neonatal jaundice, including inherited G6PD deficiency, ABO incompatibility, prematurity, prolonged labor, traumatic birth and infections (1). Standard of care should include identification of at-risk neonates, and monitoring of bilirubin levels before discharge from the birthing center. Treatment involves blue-light phototherapy and in some cases exchange transfusion based on bilirubin nomograms adjusted by gestational age and neonate’s age (2).

Newborns of low and middle income countries (LMIC) often have a late diagnosis of NH (3). Major barriers to appropriate post-natal jaundice care are the uncertainty of gestational age, the lack of diagnostic tools for risk factors and for bilirubin levels, and the poor access to treatment (especially for the most severe cases requiring exchange transfusion).

Total and unconjugated bilirubin can be measured by HPLC (the gold standard test) but most commonly total serum bilirubin (TSB) alone is measured by more widely available biochemistry analysers or by microbilirubinometers on a venous or capillary blood sample.

Non-invasive approaches have been developed to reduce neonatal pain (4). Transcutaneous bilirubinometers (TcB) are non-invasive screening tools that can detect subcutaneous concentrations of bilirubin by colorimetry from the forehead or sternum and inform need of confirmatory blood testing for diagnosis of NH and treatment. TcB have been increasingly used worldwide with varying reported performance according to age of the neonate, colour of the skin and levels of TSB. In Asia, validation studies for TcB have been carried out in India, Thailand, Malaysia, Japan, Mongolia and China (5-10) showing promising results.

Bilirubin levels measured non-invasively in cord blood have been tested for prediction of NH developing during the first 72 hours of life (11-13) and might be used as a very early risk predictor of NH (14) with the obvious advantage of not requiring an invasive heel-prick.

In this study, performance of TcB was assessed in paired non-invasive transcutaneous measures against TSB detected in capillary blood samples in a population of neonates of Karen and Burman ethnicity born in SMRU clinics along the Thailand-Myanmar border. Non-invasive cord blood bilirubin concentrations were analysed as predictors of NH in the first week of life.

## Materials and Methods

The study was conducted in SMRU clinics situated along the Thailand-Myanmar border (Thailand) where free antenatal care and birthing services are provided for migrant women of predominantly Karen and Burman ethnicities. Pregnant women attending SMRU clinics at Wang Pha (WPA) and Maw Ker Thai (MKT) were informed about the study at antenatal care visits in the 3^rd^ trimester. Informed consent was obtained before labour and eligibility of neonates was assessed immediately after delivery. Neonates born at an estimated gestational age (EGA) by ultrasound of ≥35 weeks, with no severe maternal complications at delivery and no severe neonatal illness, were included.

Two milliliters of blood were collected into EDTA from the umbilical cord and an aliquot was used for total serum bilirubin measurement. After birth, capillary blood TSB test was performed at routine pre-discharge check (at 48±12h of life) and, whenever clinically indicated, up to 1 week of life. Indication for starting phototherapy treatment followed the recommendations of the UK NICE guidelines (2) based on capillary blood TSB testing and using EGA and age adjusted nomograms. TcB was performed whenever possible, e.g. if the neonate was not crying and the TcB device was available; TcB tests were always performed shortly before blood sampling for TSB (maximum 5 minutes), and TcB results were available immediately (blinded to TSB results). Follow-up in the study was considered completed for the participant if a neonate developed NH or was admitted to the Special Care Baby Unit (SCBU). TSB measurement continued during and after phototherapy for clinical purposes only while collection of TcB measure stopped.

TSB in cord blood and capillary blood was measured by clinical laboratory technicians. An aliquot of blood was transferred into a heparinized capillary tube (70uL), centrifuged at 10,000 rpm for 3 minutes, and analysed by dual-wavelength BR-501 microbilirubinometer (Apel Co. Ldt, Japan) according to manufacturer’s instructions. Quality Controls for the microbilirubinometer were run daily, calibration was carried out weekly and external maintenance was performed yearly. The BR-501 bilirubinometer had ± 5% accuracy within the measurement range (0–30 mg/dl), as reported by the manufacturer.

TcB measures were done using a Jaundice Meter JM-105 (Dräger, Germany). The device was used in MKT clinic in 2020 and in WPA clinic from 2021 to 2023. Manufacturer’s instructions were followed to perform the daily QC procedure. Bilirubin concentrations were measured by placing the device on the neonate’s mid-sternum for 3 consecutive readings whose automatic average was used as final measure. Personnel collecting blood samples and measuring TcB were locally trained skilled birth attendants who follow a standardized SMRU curriculum (15). Training for use of the TcB was carried out twice at each clinic before the start of the study; full SOP and simplified visual aid were available during the study.

### Sample size

For the diagnostic performance of TcB compared to TSB reference test, the sample size was calculated considering estimation of the limits of agreement in Bland-Altman plot, whereby a minimum of 350 individuals is needed to obtain 95% CI of the limits of agreement within 0.2 standard deviation (SD) of the difference (16).

### Statistical analyses

Mean and SD were reported for continuous variables. Categorical variables were compared by χ^2^ test and analysis of variance.

Bland-Altman plot was used to inspect correspondence between TSB in capillary blood and TcB; intraclass correlation coefficient (ICC) was calculated for assessment of absolute agreement between paired capillary TSB and TcB values using a two-way mixed effects model. Diagnostic performance of TcB (sensitivity, specificity, positive predictive value and negative predictive value) was calculated for the following capillary TSB thresholds: ≥100 umol/L, ≥150 umol/L, then 25 umol/L increments until ≥300 umol/L.

Area under the curve (AUC) of the Receiver Operating Characteristic (ROC) curve (17) was calculated for the whole cohort, only for neonates followed-up at least for 48 hours, and at different cord blood TSB thresholds to analyse risk of NH requiring treatment. For this analysis, neonates gestational age was categorised as <38 and ≥38 weeks (18).

Data were analysed using SPSS V.27 and statistical significance was assessed at the 5% level.

### Patient and public involvement

At the outset of the study, the research team engaged the local population through the Tak Province Community Advisory Board, Thailand. This group is comprised of community leaders who were asked to advise on study design, process and outcomes of interest, and subsequently approved the study.

## Results

Between April 2020 and May 2023, a total of 742 neonates born at SMRU facilities (477 in MKT and 265 in WPA) entered the study and provided cord blood samples of which 738 had a valid TSB measurement (Figure 1). Mean (SD) EGA in the cohort was 39.3 (1.0) weeks, 49 neonates (6.6%) were born with EGA 35 to <38 weeks and 49.6% of neonates were female.

**Figure 1.**
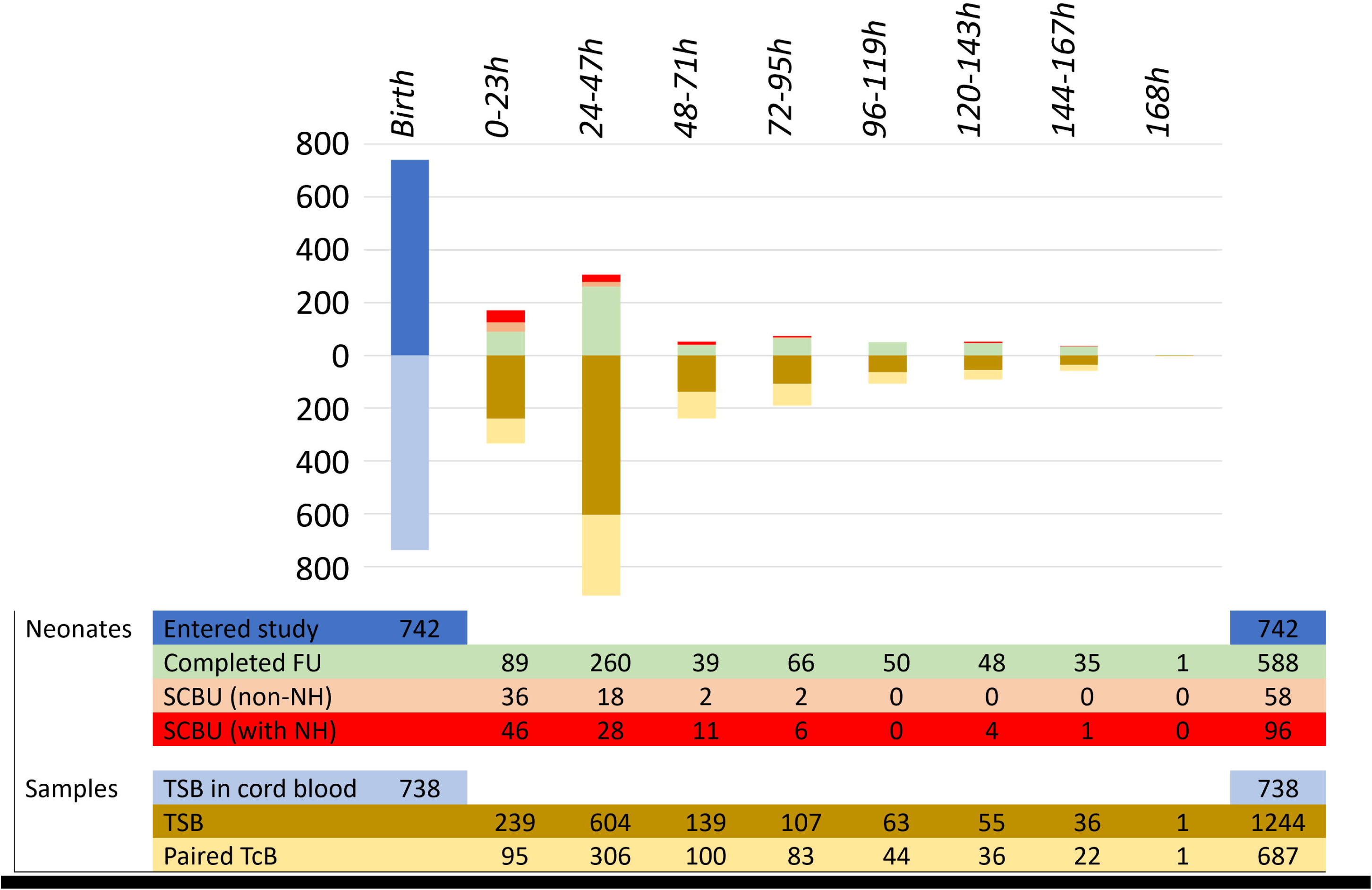
Participants and samples

Within the first 48 hours, 47 neonates completed the study without a capillary TSB or TcB measurement; 43 neonates were admitted to SCBU, 2 were referred to other hospitals and 2 were discharged. In the following 120 hours, the remaining 695 neonates provided 1 to 9 additional capillary blood samples for analysis of TSB at a mean age of 47h for a total of 1244 analyzable samples. Over half of the capillary TSB data point (687 from 307 neonates, 55.2%) had a paired TcB value.

During the first 7 days of life, admission to SCBU was recorded in a total of 154 neonates (43 within the first 48 hours and 111 up to 158 h of life) mostly for early onset of neonatal sepsis and/or NH; phototherapy was recorded in 96 neonates (12.8%), three of whom were treated twice. Around half of neonates (358/742) had their last bilirubin checked at the second or third day of life before discharge from the clinic; around 30% had TSB done at follow-up between 4 and 7 days of life.

### Transcutaneous Bilirubin *versus* Capillary Total Serum Bilirubin during the first week of life

A total of 1250 capillary TSB tests results were collected (718 in MKT and 532 in WPA); six data points were excluded from analysis (Supplementary Material, Figure S1) for a total of 1244 usable samples.

A total of 687 paired TcB-capillary TSB tests from 307 neonates were available in MKT clinic (448 tests) and in WPA clinic (239 tests). Paired measures were collected between 2 and 168h of life. Mean difference (±1.96SD) in bilirubin levels between TcB and capillary TSB was 20.5 umol/L (Limits of Agreement -42.7 to 83.6 umol/L) as shown in the Bland-Altman plot in Figure 2. The analysis included multiple tests performed in the same individuals, slightly wider LoA were obtained when calculated separately for consecutive TcB-capillary TSB repeat (Table S1).

**Figure 2.**
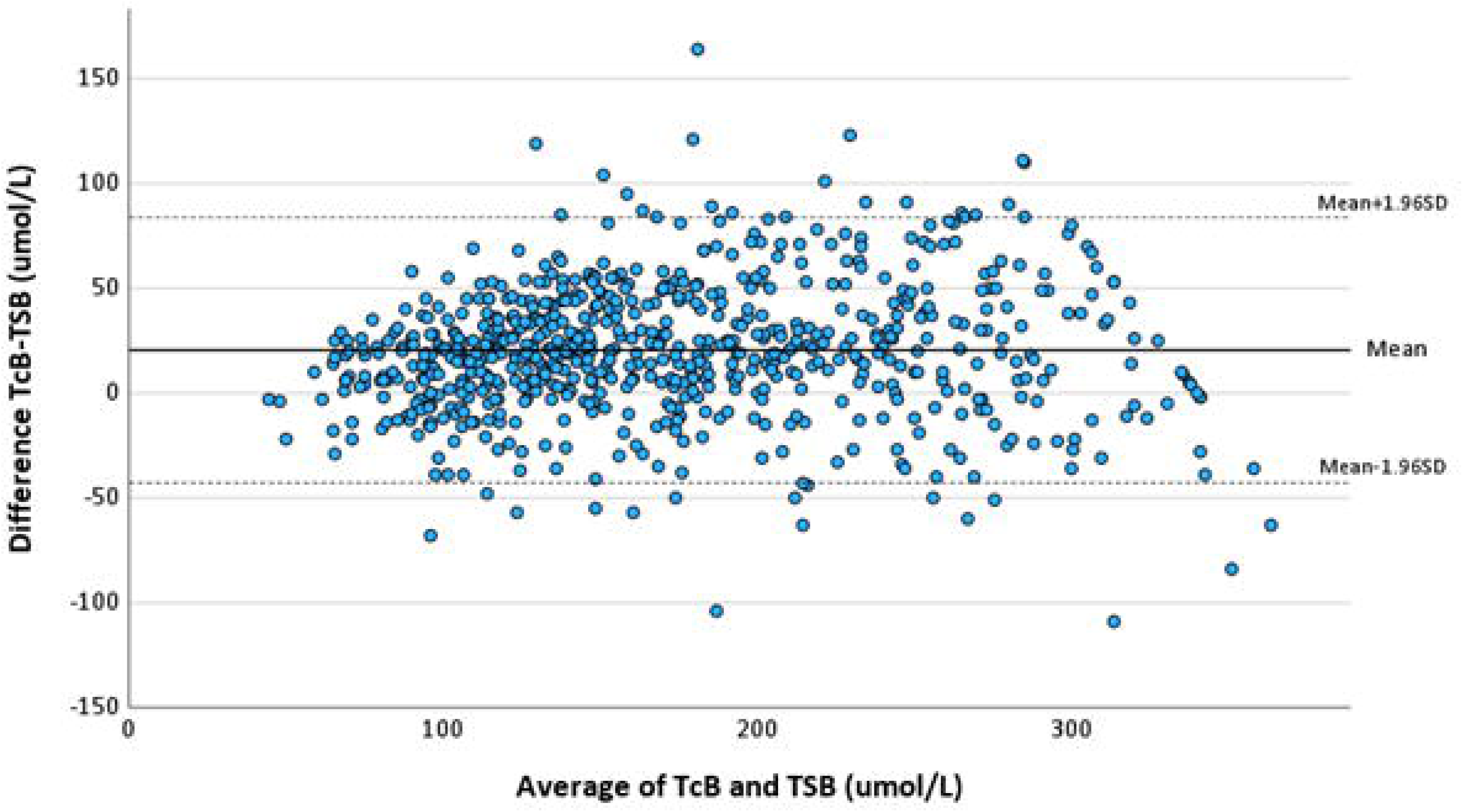
Bland-Altman plot of bilirubin concentration assessed by TcB as compared to TSB.

Intraclass correlation coefficient (ICC) showed excellent agreement between paired TcB-capillary TSB tests overall (0.923, 95%CI:0.818-0.958, P<0.01) and across neonates’ ages (Table 3).

**Table 3.**
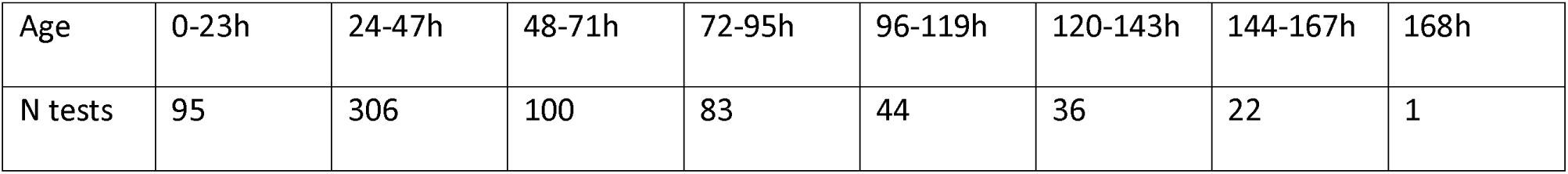
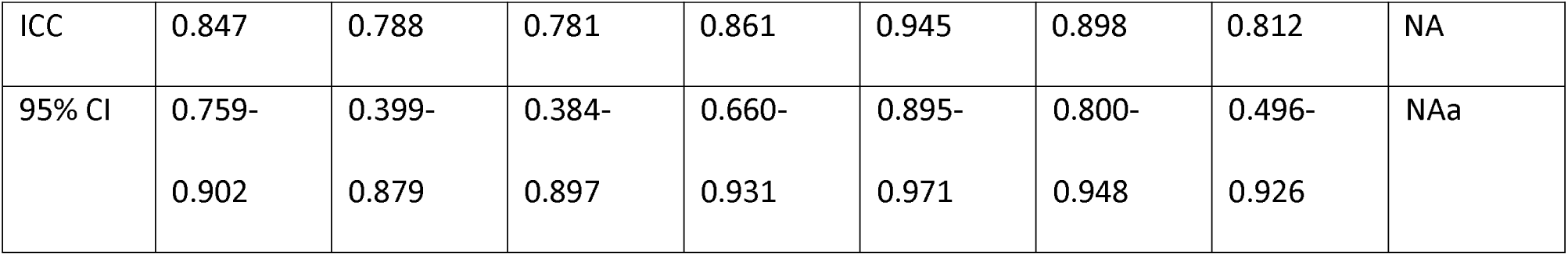
Intraclass Correlation coefficient for paired TcB-capillary TSB tests across neonates’ age.

Performance of the TcB, calculated as sensitivity and specificity to identify capillary TSB levels over different thresholds, was improved when 20umol/L were added to the reading from the TcB (Table S2), especially at higher thresholds.

Results showed a sensitivity over 98% at all the thresholds analysed while specificity was usually lower at lower thresholds (100 to 150umol/L) as compared to higher thresholds (250 to 300 umol/L) and declined slightly over time. TcB overall performance, and performance by clinic when using the adjusted (TcB+20) values, and performance over time are reported in Table S3 and S4.

In a small sample of neonates (n=26), five or more paired longitudinal TcB-capillaryTSB tests were available and their consistent time-course of bilirubin concentrations is shown in Figure S2.

### Total Serum Bilirubin in cord blood for prediction of NH

Among all 738 neonates with cord blood TSB analysable data, mean (SD) cord blood TSB levels were 30.8 (11.7) umol/L (Figure S3). They were higher in neonates born with EGA 35 to <38 weeks (34.4umol/L) compared to neonates born with EGA≥38 weeks (30.5umol/L; P<0.01).

Statistically higher mean (SD) cord TSB [41.0 (17.6) umol/L] was observed in the 96 neonates who required phototherapy during the first week of life as compared to neonates who did not [29.1 (9.5) umol/L; P<0.001]. AUC of ROC curve was 0.771 (95%CI: 0.722-0.819) for the whole cohort and 0.738 (95% CI: 0.634-0.842) when including only neonates followed up for at least 48 hours (n=263), Figure 3. ROC analysis could not identify thresholds of cord blood TSB that were predictive of hyperbilirubinaemia and need of phototherapy during the follow-up period (Table S5).

**Figure 3.**
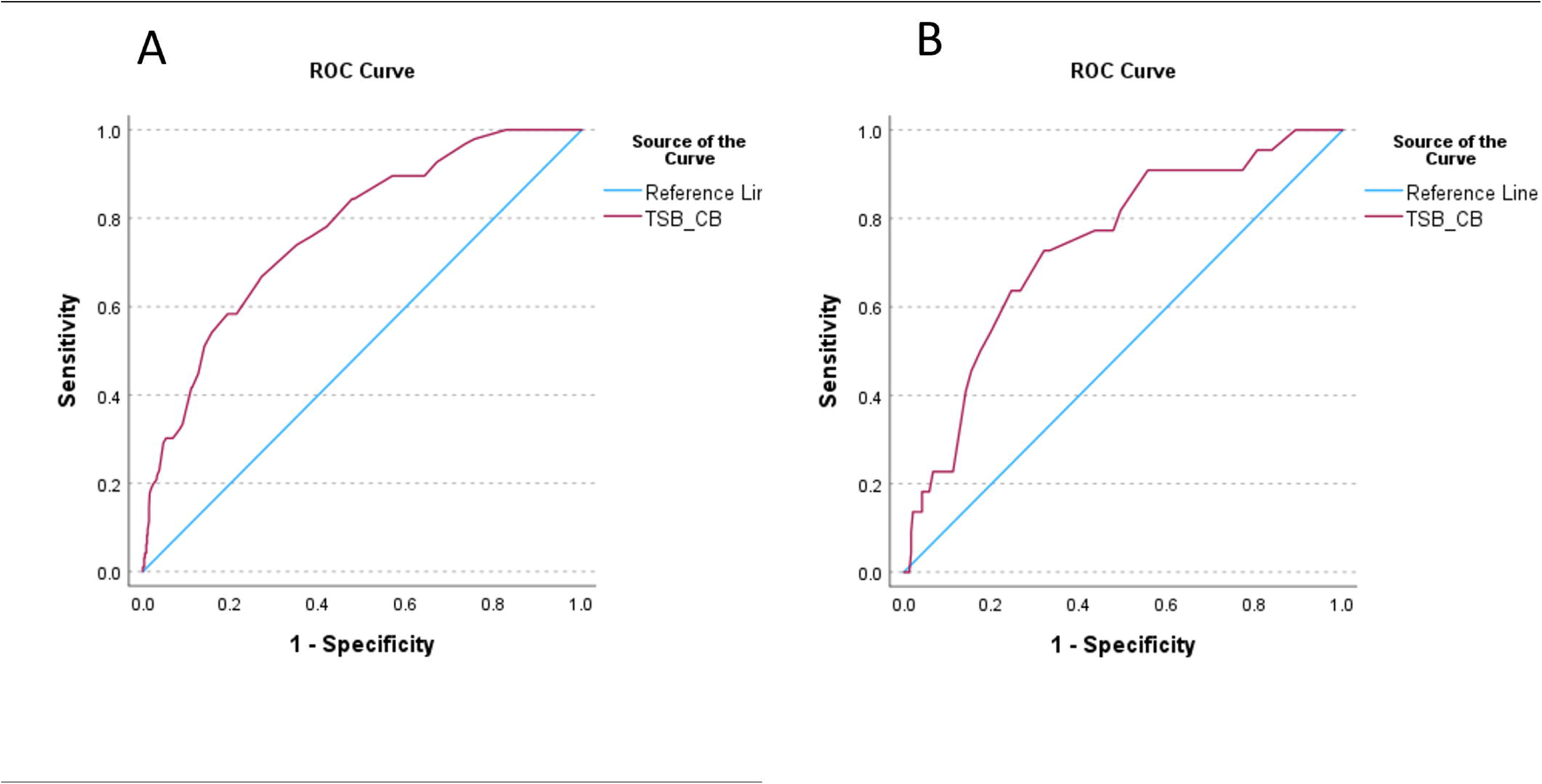
ROC curves for cord-blood bilirubin levels predicting NH (A: whole cohort (n=738); B: only neonates followed-up for at least 48h (n=263)

## Discussion

The current study evaluated non-invasive screening tools to improve clinical care of neonates in a low resource setting.

Transcutaneous bilirubin assessments are routinely used in many Western Countries and in hospital settings of some tropical countries. Their reliability in populations with more pigmented skin has not been tested widely (19-21) and was evaluated against capillary TSB levels in neonates of Karen and Burman ethnicity here for the first time. Results from this study indicate that TcB values, with an appropriate simple correction, can be used reliably against TSB nomograms to identify neonates who need confirmatory laboratory blood tests to detect clinically significant bilirubin levels. Measurements taken by multiple different users over 3 years (March 2020 to May 2023) in neonates born in two different clinics, with different EGA and aged 2 hours up to 6 days of life, showed very good performance of the TcB in real-life conditions.

Mean difference against capillary TSB was in line with accuracy data provided by the manufacturer (± 25.5 umol/L in neonates >35 weeks gestation) and ICC was excellent. Performance at different TSB thresholds (22) showed very good sensitivity, indicating that screening with TcB can identify correctly neonates with higher TSB values who would need a confirmatory blood TSB test. Even with the low specificity observed especially at 100 or 150umol/L TSB thresholds (which would translate to performing unnecessary blood tests in neonates with true levels of bilirubin lower than those thresholds), the use of TcB would decrease the total number of invasive blood testing required as compared to current practice. Routine use of TcB could also promote more frequent non-invasive assessment of bilirubin concentrations to identify dangerously steep raises in bilirubin levels associated with a more severe course of NH (2, 23). Use of TcB might be extended to more remote settings as the Drager JM-105 requires minimal maintenance and no consumables. However, use of devices in field conditions and performance of long-term use in extreme temperature/humidity conditions would have to be tested (especially for battery life). The price (around 7,000 USD) represents an initial insurmountable investment for most constrained-resource settings, but other transcutaneous devices might be as accurate and more affordable (24). Other cheaper screening tools have been tested, such as icterometers (25, 26) and phone-based apps (27-29), and when properly evaluated could be used by health workers at birthing centers and in remote settings, and even by parents (30) after discharge from the hospital.

This analysis of bilirubin concentrations in cord blood found no meaningful thresholds for identification of neonates at risk of NH, in all neonates or in the sub-group of term neonates. These findings contrast with previously published results (11, 31) but confirm the recommendation from NICE Guidelines not to use cord blood bilirubin concentrations to predict hyperbilirubinaemia (2).

Limitations of the study include not testing neonates during phototherapy where higher TSB values are observed; this indication is now included in the test specifications (recently reviewed by Ten Kate et al (24)). Usability of the device by locally trained clinical staff was not formally assessed; however, during discussions with the midwives involved, the device was deemed easy to use. Finally, the device was not serviced every 12 months as per manufacturer’s instructions due to COVID-19 pandemic and this might have had an impact on performance over time.

## Conclusions

Most death and disability from NH occur in LMIC, and this loss of healthy life-years is preventable with proper identification and treatment. Together with improved education, there are still measures that can be implemented to improve care for neonatal jaundice in LIMC (32). Better and affordable diagnostics are needed globally (33, 34) and especially in low-resource and conflict areas where clinical care is often dysfunctional. While screening of bilirubin in cord blood does not appear to provide benefit in identifying most at-risk neonates, TcB can be used reliably as a screening tool to inform need of confirmatory blood testing, supporting more frequent pain-free bilirubin checks and informing better clinical care for neonates in LMIC.

## Ethics

This study involves human participants and was approved by the Oxford Tropical Research Ethics Committee, UK (OxTREC 532-19), the Mahidol University Faculty of Tropical Medicine Ethics Committee, Thailand (TMEC 19-048, MUTM 2019-080-02) and the Tak Province Border Community Ethics Advisory Board (TCAB201904). Written informed consent was obtained from literate mothers and a thumbprint was obtained in the presence of a literate witness for illiterate mothers.

## Data availability statement

De-identified participant data are available from the Mahidol Oxford Tropical Medicine Data Access Committee upon request from this link: https://www.tropmedres.ac/units/moru-bangkok/bioethics-engagement/datasharing.

## Acknowledgments

The authors wish to thank all the mothers for their collaboration and understanding; the study would not have been possible without the hard work and dedication of all SMRU workers involved, especially during such a difficult time of political unrest and the COVID-19 pandemic.

## Contributions

Substantial contributions to the conception or design of the work—GB, FN, VIC and RM. Acquisition, analysis or interpretation of data for the work—GB, MEG, EW, GG, PKM, LA, NSW, SW, BH, FN, VIC and RM. Drafting the work—GB. Revising the work critically for important intellectual content— MEG, EW, GG, PKM, LA, NSW, SW, BH, FN, VIC and RM. Final approval of the version to be published—all authors.

