## Supplementary materials for "Non-invasive detection of bilirubin concentrations during the first week of life in a low-resource setting along the Thailand-Myanmar border"

### Supplementary Material

**Excluded TcB-TSB paired data points**

A total of 6 data points were excluded (circled in blue) from analyses based on the longitudinal data trends as shown in Figure S3.

From the top pane, in the first participant the TSB#4 was probably a reporting mistake; in the second participant, samples #4 and #5 were collected at 5am and 8:40am, we suspect that TSB#4 and TcB#5 could be reporting mistakes. In the third participant, TSB#1 was 35 umol/L with hct=73% while TSB#2 (collected 2hrs later) was 106umol/L with hct=62% therefore we suspected a problem in the sample used for TSB#1 test. In the fourth participant, the two TcB records were exchanged in the logbook but we could not confirm the correct result.

**Figure S1. Excluded paired TSB-TcB data points**

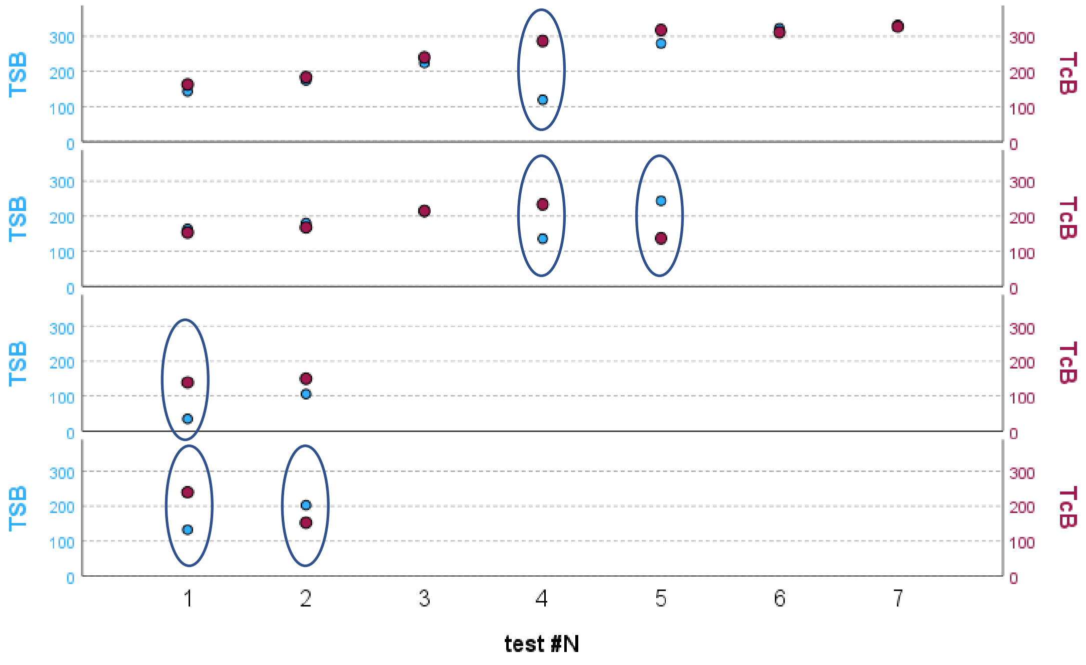

**Table S1. Mean difference TcB-TSB for consecutive TSB tests**

| TSB | N | Mean Difference TcB-TSB | Lower LoA | Upper LoA |
|---|---|---|---|---|
| 1st | 296 | 19.8 | -36.4 | 76.0 |
| 2nd | 211 | 25.6 | -40.0 | 91.3 |
| 3rd | 87 | 19.5 | -48.0 | 87.0 |
| 4th | 43 | 17.8 | -59.9 | 95.5 |
| 5th | 26 | 15.8 | -42.3 | 74.0 |
| 6th | 13 | 4.5 | -55.2 | 64.1 |
| 7th | 5 | -14.0 | -53.7 | 25.7 |
| 8th | 3 | -34.0 | -126.7 | 58.7 |
| 9th | 1 | 0.0 | | |
| Total | 685 | 20.5 | -42.7 | 83.6 |

**Table S2. Sensitivity and specificity of TcB without and with correction at different bilirubin thresholds in all samples**

| TSB threshold | Sensitivity TcB without correction | Specificity TcB without correction | Sensitivity TcB with correction | Specificity TcB with correction |
|---|---|---|---|---|
| ≥100 | 0.97 | 0.50 | 1.00 | 0.20 |
| ≥150 | 0.96 | 0.69 | 0.99 | 0.48 |
| ≥175 | 0.94 | 0.78 | 0.98 | 0.64 |
| ≥200 | 0.95 | 0.84 | 0.99 | 0.73 |
| ≥225 | 0.95 | 0.87 | 0.96 | 0.79 |
| ≥250 | 0.88 | 0.88 | 0.98 | 0.83 |
| ≥275 | 0.78 | 0.93 | 0.88 | 0.87 |
| ≥300 | 0.72 | 0.95 | 0.90 | 0.91 |

**Table S3. Performance of the TcB+20 at different bilirubin thresholds by clinic**

| All clinics | Threshold (umol/L) | N (TcB>= threshold) | Sensitivity | Specificity | PPV | NPV |
|---|---|---|---|---|---|---|
| | ≥100 | 661 | 1.00 | 0.20 | 0.86 | 0.92 |
| | ≥150 | 517 | 0.99 | 0.48 | 0.65 | 0.97 |
| | ≥175 | 413 | 0.98 | 0.64 | 0.63 | 0.98 |
| | ≥200 | 328 | 0.99 | 0.73 | 0.60 | 1.00 |
| | ≥225 | 250 | 0.96 | 0.79 | 0.55 | 0.99 |
| | ≥250 | 188 | 0.98 | 0.83 | 0.46 | 1.00 |
| | ≥275 | 134 | 0.88 | 0.87 | 0.38 | 0.99 |
| | ≥300 | 84 | 0.90 | 0.91 | 0.31 | 1.00 |

| | | | | | | |
|---|---|---|---|---|---|---|
| WPA | | | | | | |
| | ≥100 | 235 | 1.00 | 0.09 | 0.83 | 1.00 |
| | ≥150 | 195 | 0.99 | 0.32 | 0.52 | 0.98 |
| | ≥175 | 156 | 0.99 | 0.51 | 0.49 | 0.99 |
| | ≥200 | 117 | 1.00 | 0.65 | 0.43 | 1.00 |
| | ≥225 | 90 | 1.00 | 0.71 | 0.33 | 1.00 |
| | ≥250 | 70 | 1.00 | 0.75 | 0.21 | 1.00 |
| | ≥275 | 50 | 1.00 | 0.81 | 0.12 | 1.00 |
| | ≥300 | 30 | NA | NA | NA | NA |
| MKT | | | | | | |
| | ≥100 | 426 | 0.99 | 0.27 | 0.87 | 0.91 |
| | ≥150 | 322 | 0.98 | 0.58 | 0.72 | 0.97 |
| | ≥175 | 257 | 0.97 | 0.72 | 0.72 | 0.97 |
| | ≥200 | 211 | 0.99 | 0.79 | 0.70 | 1.00 |
| | ≥225 | 160 | 0.96 | 0.84 | 0.67 | 0.98 |
| | ≥250 | 118 | 0.97 | 0.88 | 0.61 | 0.99 |
| | ≥275 | 84 | 0.87 | 0.90 | 0.54 | 0.98 |
| | ≥300 | 54 | 0.90 | 0.93 | 0.48 | 0.99 |

PPV= positive predictive value; NPV=negative predictive value

**Table S4. Performance of corrected TcB thresholds over time**

| | MKT clinic | | | WPA clinic | | | | | | | | |
|---|---|---|---|---|---|---|---|---|---|---|---|---|
| | 2020 | | | 2021 | | | 2022 | | | 2023 | | |
| ALL | N (TcB>= threshold) | se | sp | N (TcB>= threshold) | se | sp | N (TcB>= threshold) | se | sp | N (TcB>= threshold) | se | sp |
| 150 | 321 | 0.98 | 0.58 | 8 | 1 | 0.33 | 147 | 0.99 | 0.32 | 39 | 1 | 0.26 |
| 200 | 210 | 0.99 | 0.79 | 5 | 1 | 0.63 | 85 | 1 | 0.65 | 26 | 1 | 0.62 |
| 250 | 117 | 0.97 | 0.88 | 2 | 1 | 0.89 | 49 | 1 | 0.77 | 18 | 1 | 0.68 |
| 300 | 53 | 0.89 | 0.93 | 2 | NA | NA | 18 | NA | NA | 10 | NA | NA |

**Figure S2. Examples of longitudinal course of repeated paired TcB-capillary TSB tests in different neonates (1 neonate per pane).**

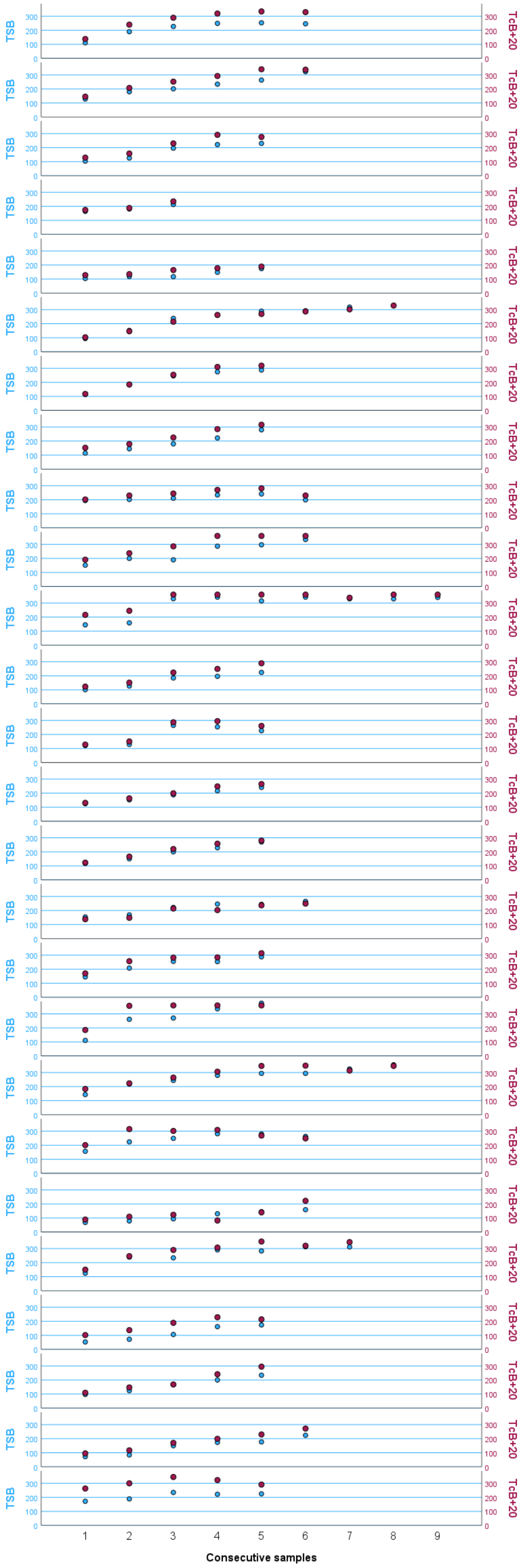

**Figure S2. Distribution of TSB in cord blood by clinical site**

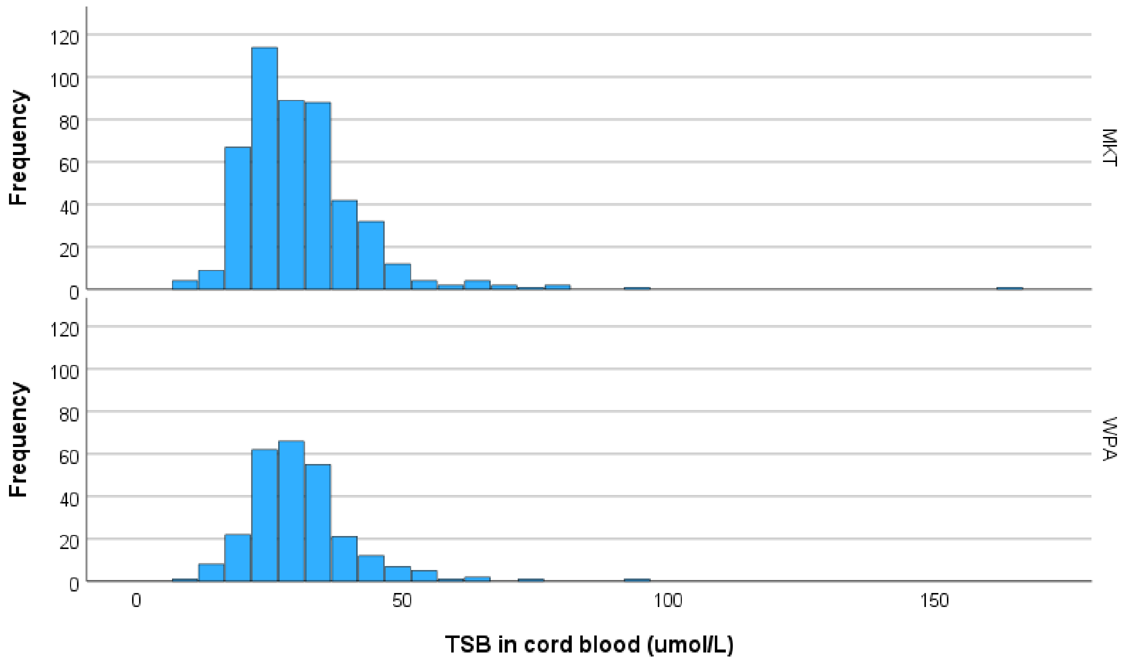


**Table S5. Diagnostic performance of cord blood TSB thresholds for identification of NH**

| Threshold | Population | Reference | Sensitivity | Specificity |
|---|---|---|---|---|
| >32.0umol/L | All neonates | (1) | 67.4% | 73.1% |
| >32.0umol/L | Only EGA≥38 weeks | (1) | 77.3% | 72.5% |
| >35.0umol/L | Only EGA≥38 weeks | (2) | 71.2% | 80.3% |
